# MIMOSA: A resource consisting of improved methylome imputation models increases power to identify DNA methylation-phenotype associations

**DOI:** 10.1101/2023.03.20.23287418

**Authors:** Hunter J. Melton, Zichen Zhang, Hong-Wen Deng, Lang Wu, Chong Wu

## Abstract

Although DNA methylation has been implicated in the pathogenesis of numerous complex diseases, the exact methylation sites that play key roles in these processes remain elusive. One strategy to identify putative causal CpG sites and enhance disease etiology understanding is to conduct methylome-wide association studies (MWASs), in which predicted DNA methylation that is associated with complex diseases can be identified.However, current MWAS models are primarily trained by using the data from single studies, thereby limiting the methylation prediction accuracy and the power of subsequent association studies. Here, we introduce a new resource, MWAS Imputing Methylome Obliging Summary-level mQTLs and Associated LD matrices (MIMOSA), a set of models that substantially improve the prediction accuracy of DNA methylation and subsequent MWAS power through the use of a large, summary-level mQTL dataset provided by the Genetics of DNA Methylation Consortium (GoDMC). With the analyses of GWAS (genome-wide association study) summary statistics for 28 complex traits and diseases, we demonstrate that MIMOSA considerably increases the accuracy of DNA methylation prediction in whole blood, crafts fruitful prediction models for low heritability CpG sites, and determines markedly more CpG site-phenotype associations than preceding methods. Finally, we use MIMOSA to conduct a case study in high cholesterol, pinpointing 146 putatively causal CpG sites.

## 1. Introduction

DNA methylation (DNAm), the epigenetic mechanism of adding methyl groups (CH_3_) to the genome, serves an essential role in the regulation of gene expression [1]. Inter-individual variation in DNAm has been shown to be linked to the etiology of many complex traits through conventional epidemiological studies. However, its impact on disease development is not yet well-characterized, primarily due to several common issues encountered in observational studies, including selection bias, unmeasured confounding factors, and reverse causation. One strategy to address such limitations is employing methylome-wide association studies (MWASs) [2–4]. Similar to transcriptome-wide association studies (TWASs) [5–8], MWAS employs an instrumental variable regression setup involving two steps. First, MWAS builds DNA methylation prediction models for a set of CpG sites using a methylation reference panel where both genetic and DNA methylation data are available. Second, MWAS tests the association between predicted DNA methylation levels and traits of interest.

While appealing, current MWASs, to our knowledge, rely on individual-level reference panels to build DNAm prediction models, thus limiting statistical power due to the limited sample sizes of these reference panels. To ameliorate this limitation, we introduce the MWAS Imputing Methylome Obliging Summary-Level mQTLs and Associated LD matrices (MIMOSA), a novel and comprehensive MWAS resource that improves DNAm prediction accuracy beyond existing models [2, 3] and can be used to boost MWAS power. Inspired by a new TWAS method SUMMIT [8] that builds expression prediction models using summary-level expression quantitative trait loci (eQTLs) data, we build DNAm imputation models using summary-level methylation quantitative trait loci (mQTLs) data in whole blood (*n* = 27, 750 individuals) from the Genetics of DNA Methylation Consortium (GoDMC) [9]. Briefly, the GoDMC mQTL associations are based on samples from 27,750 participants of European ancestry hailing from 36 independent studies. The DNAm was profiled in blood with Illumina HumanMethylation450 BeadChips or EPIC arrays in at least 100 participants. Likely due to limitations of available sample sizes of individual-level methylation reference panels, only imputation models with *R*^2^ *>* 0.01 are included in downstream association analyses [2, 4], as methylation at CpG sites with lower heritability was not previously able to be accurately modeled. In contrast, MIMOSA includes imputation models with 0.005 *< R*^2^ ≤ 0.01 (which is recommended by SUMMIT), in an effort to test CpG sites with low heritability that have been largely ignored by existing MWAS models.

Through analyses of GWAS summary statistics for 28 complex traits and diseases, we demonstrate that MIMOSA successfully and substantially increases the accuracy of DNAm imputation in blood and hence subsequently identifies markedly more CpG site-trait associations than existing methods. We provide a case study in high cholesterol and a database of our newly-built DNA methylation imputation models (MIMOSA). Our association results from these GWAS are available as a resource to the community.

## 2. Results

### 2.1. MIMOSA summary

By leveraging summary-level mQTL data from GoDMC, we built MIMOSA, a set of comprehensive DNAm prediction models. Specifically, following SUMMIT [8], we trained DNAm prediction models for each of 123,375 CpG sites using five penalized regression methods: the elastic net [10], MNet [11], the smoothly clipped absolute deviation (SCAD) [12], the minimax concave penalty (MCP) [13], and LASSO [14]. We draw summary-level mQTL data in blood (*n* = 27, 750 individuals) from the GoDMC [9], which is based on Illumina HumanMethylation BeadChips and Epic arrays. Then, the models are tuned and validated using independent datasets from the Framingham Heart Study (FHS) [15]. In particular, we used FHS subcohort 1 (*n* = 797) for tuning and the independent FHS subcohort 2 (*n* = 798) for validation, and correlated samples with an estimated identity by descent (IBD) sharing 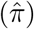 of more than 0.2 were excluded in the FHS datasets. The FHS datasets encompass mQTLs in whole blood for 370,785 CpG sites (that passed standard quality control steps as detailed in [4]), 123,375 of which were included in the summary-level training data from GoDMC. Thus, we built DNAm prediction models for these 123,375 CpG sites, as our method requires both the training data from GoDMC and the tuning data from the FHS. Lastly, models trained by MIMOSA could be used in a standard MWAS: we demonstrated this by testing the association between imputed methylation and phenotype for each methylation imputation model with satisfactory performance (*R*^2^ *>* 0.005, positive correlation), resulting in a set of CpG site-trait associations. The model framework is shown in Figure 1.

**Figure 1.**
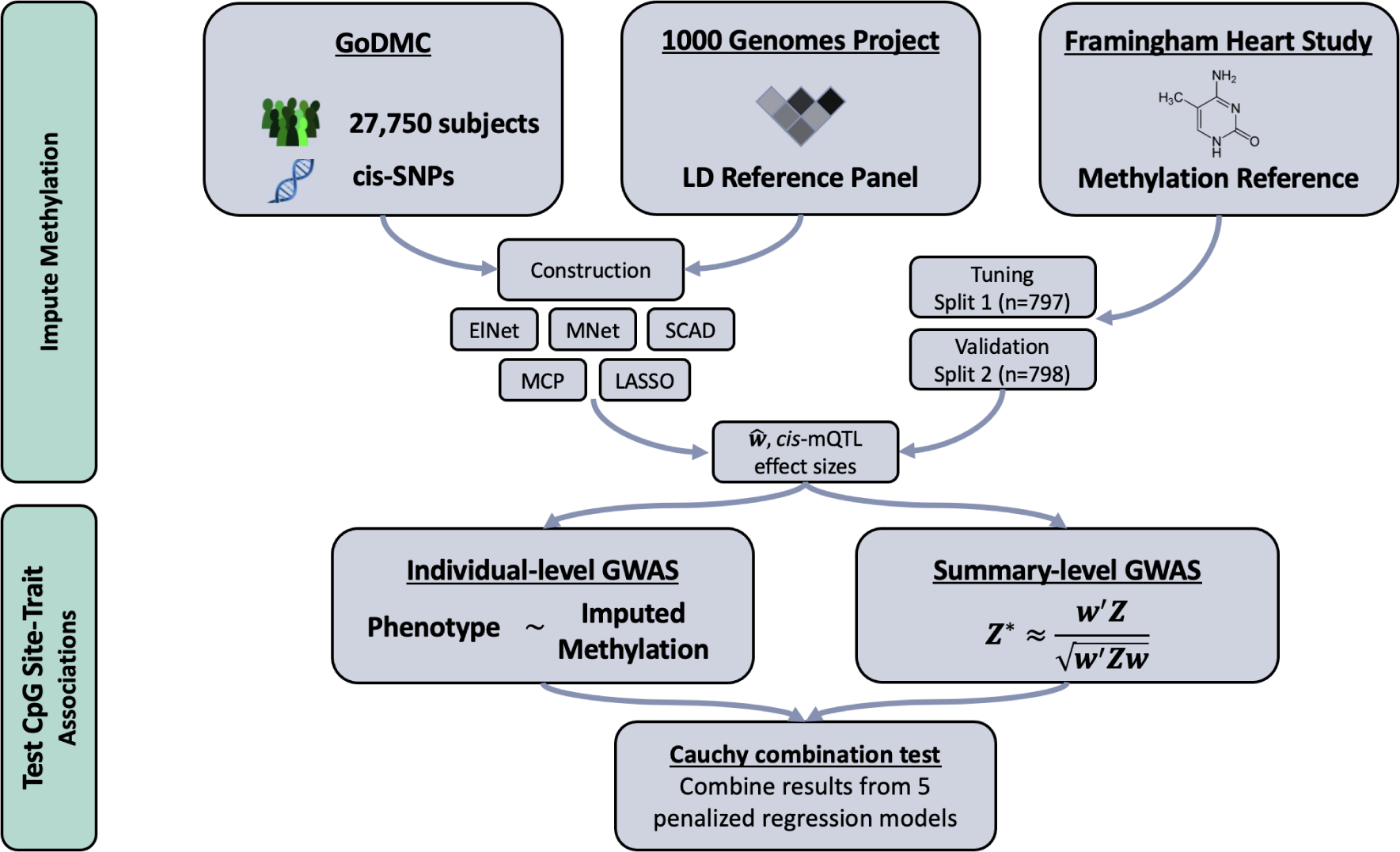
MIMOSA Workflow. MIMOSA evaluates CpG site-trait associations by: 1st) training methylome imputation models; 2nd) testing associations between imputed methylation levels and phenotype. Here, “*cis*-SNPs” referes to single nucleotide polymorphisms within 1 Mb of a particular CpG site, and “LD Reference Panel” refers to the linkage disequilibrium reference panel from the 1000 Genomes Project.

### 2.2. MIMOSA increases accuracy in methylome imputation models

We compared the imputation accuracy between MIMOSA and the models created by Baselmans et al. [2] (hereafter referred to as the Baselmans models) for whole blood using independent samples from the Framingham Heart Study (FHS). The MIMOSA models were trained on summary-level mQTL data from GoDMC [9], which is itself based on a sample size of 27,750 participants, and tuned using individual-level mQTL data (subcohort 1, *n* = 797) from the FHS [15]. In total, MIMOSA created models for 123,375 CpG sites. The Baselmans models [2] were trained on individual-level mQTL data from the BIOS Consortium [16], based on samples from 4,008 participants. We evaluated the performance of both methods using independent individual-level data (subcohort 2, *n* = 798) from the FHS [15]. To make a fair comparison, we examined the number of CpG sites with *R*^2^ *>* 0.01 and positive Pearson’s correlation for genetically predicted vs. directly measured DNAm levels for the overlapping CpG site list (i.e., CpG sites with models created by both methods). *R*^2^ is the coefficient of determination, and it refers to the proportion of variance of the methylation that can be explained by the DNA methylation prediction models.

Of the 109,722 candidate CpG sites in the overlapping CpG site list, MIMOSA built satisfactory (*R*^2^ *>* 0.01, positive correlation) imputation models for 74,283, and Baselmans for 57,223. Of the 57,223 CpG sites for which the Baselmans method created a satisfactory model, MIMOSA successfully built models for 55,131 (96.3%). Furthermore, MIMOSA yielded better imputation models for 57,637 out of 76,632 (75.2%) CpG sites with *R*^2^ *>* 0.01 and positive correlation by either model. With regard to correlation between real and predicted DNAm levels, the MIMOSA models produce a higher value than the Baselmans models for 78, 674 of the 109,722 common CpG sites (71.7%). Average correlation for the MIMOSA models was 0.20 and for the Baselmans models was 0.14. Correlation was negative for 4, 689 CpG sites (4.3%) for the MIMOSA models and 14, 251 CpG sites (13.0%) for the Baselmans models. We conducted paired, one-sided Wilcoxon signed-rank tests to compare both *R*^2^ and correlation for the full set of 109,722 CpG sites; MIMOSA significantly improved prediction accuracy overall over the Baselmans method (*p <* 2.2 × 10^−16^ for both *R*^2^ and correlation). Figure 2 demonstrates the improved imputation accuracy of MIMOSA models. It is common in TWAS or MWAS to analyze genes or CpG sites with *R*^2^ *>* 0.01 to meet the relevance assumption. In contrast, we recommend including CpG sites with *R*^2^ *>* 0.005 and positive correlation in analyses (see [8] for justifications). By using this more liberal threshold, we increased the number of testable imputation models by an additional 11,299 CpG sites.

**Figure 2.**
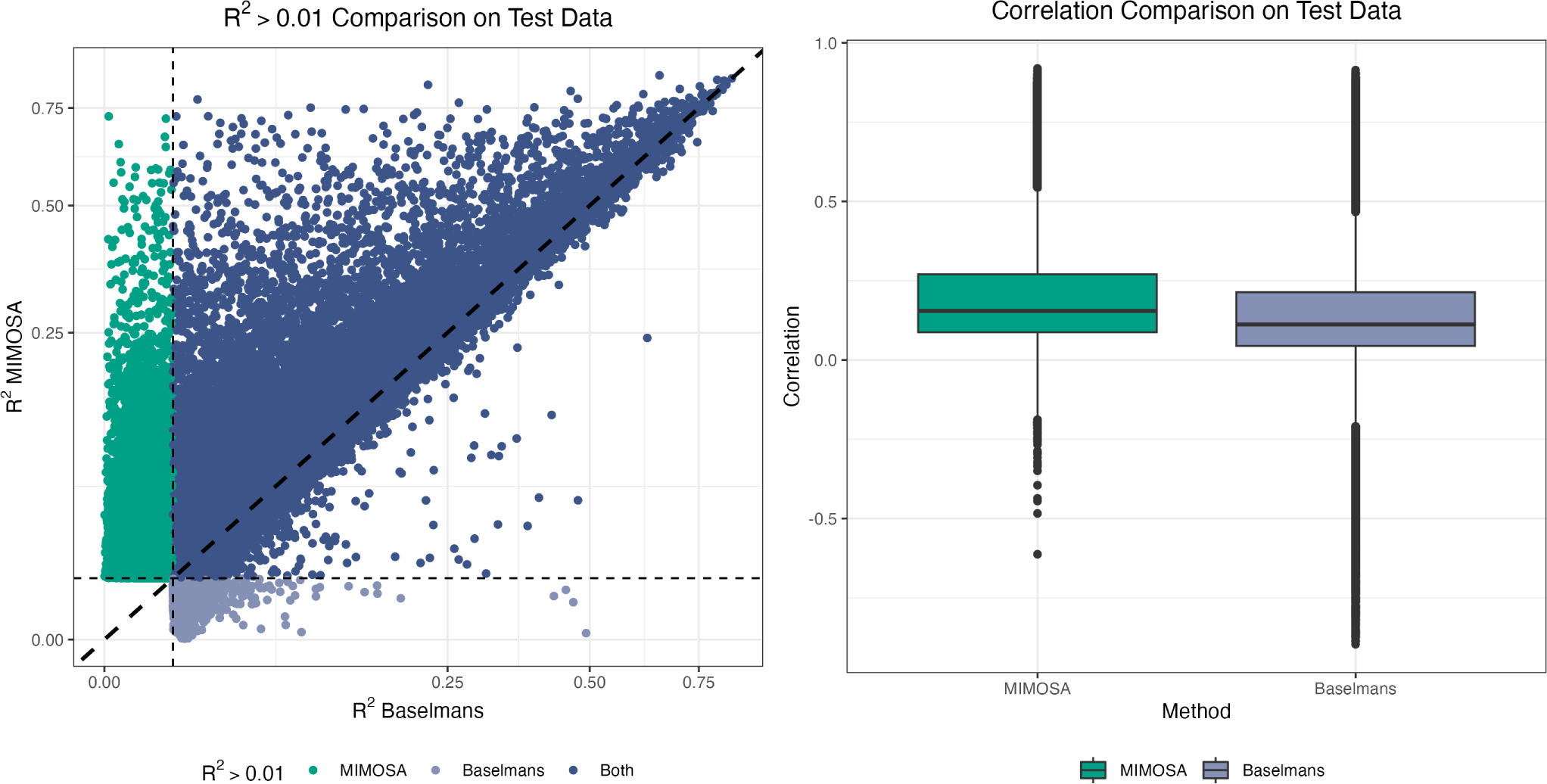
*R*^2^ Comparison. A comparison of best imputation model *R*^2^ on test data for all CpG sites for which either MIMOSA or Baselmans hit *R*^2^ *>* 0.01 threshold.

**Figure 3.**
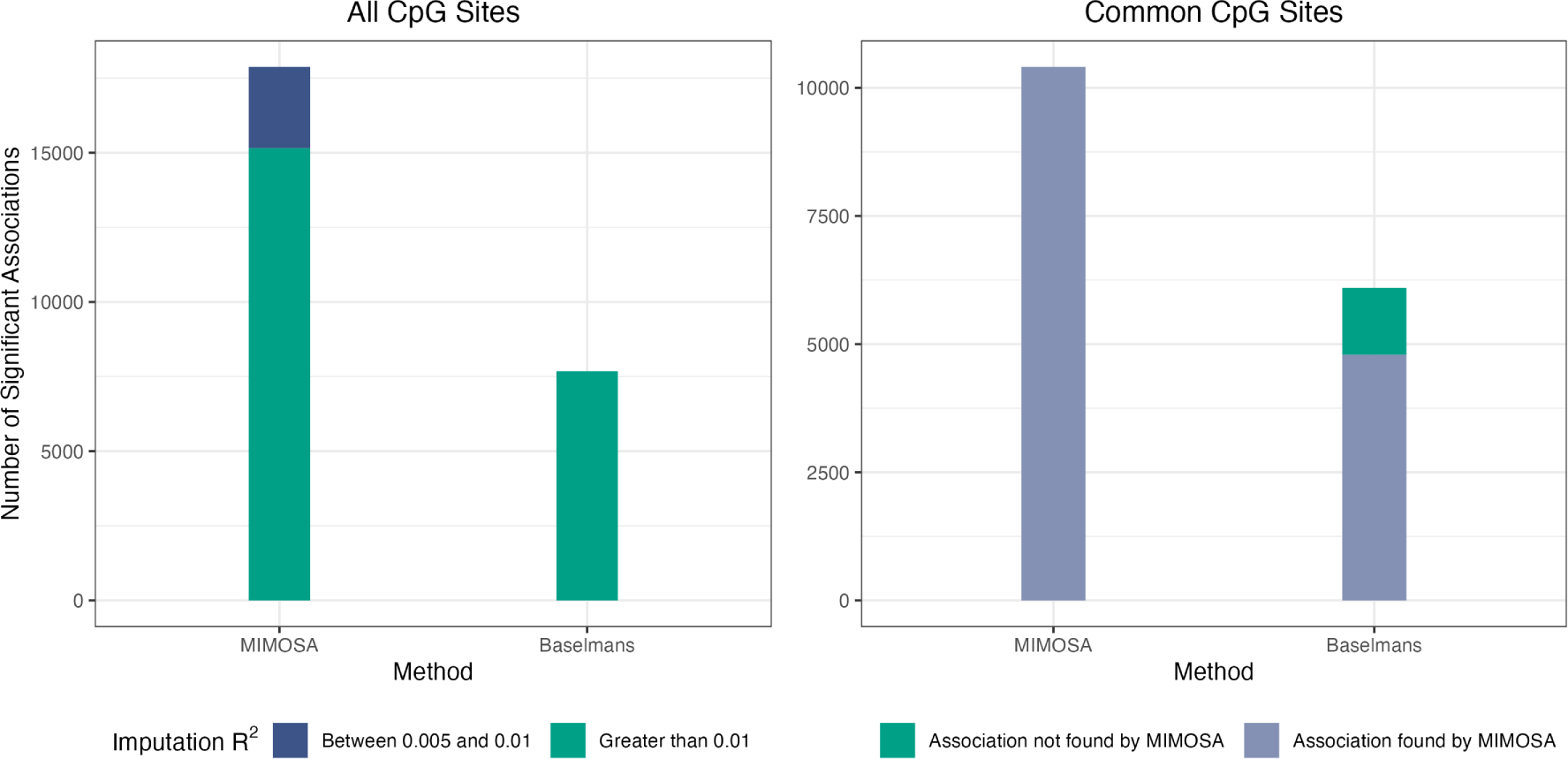
Evaluation of MIMOSA on real data. a) Number of significant associations determined by MIMOSA and Baselmans for 28 GWAS using all CpG sites. b) Number of significant associations determined by MIMOSA and Baselmans for 28 GWAS using shared CpG sites.

**Figure 4.**
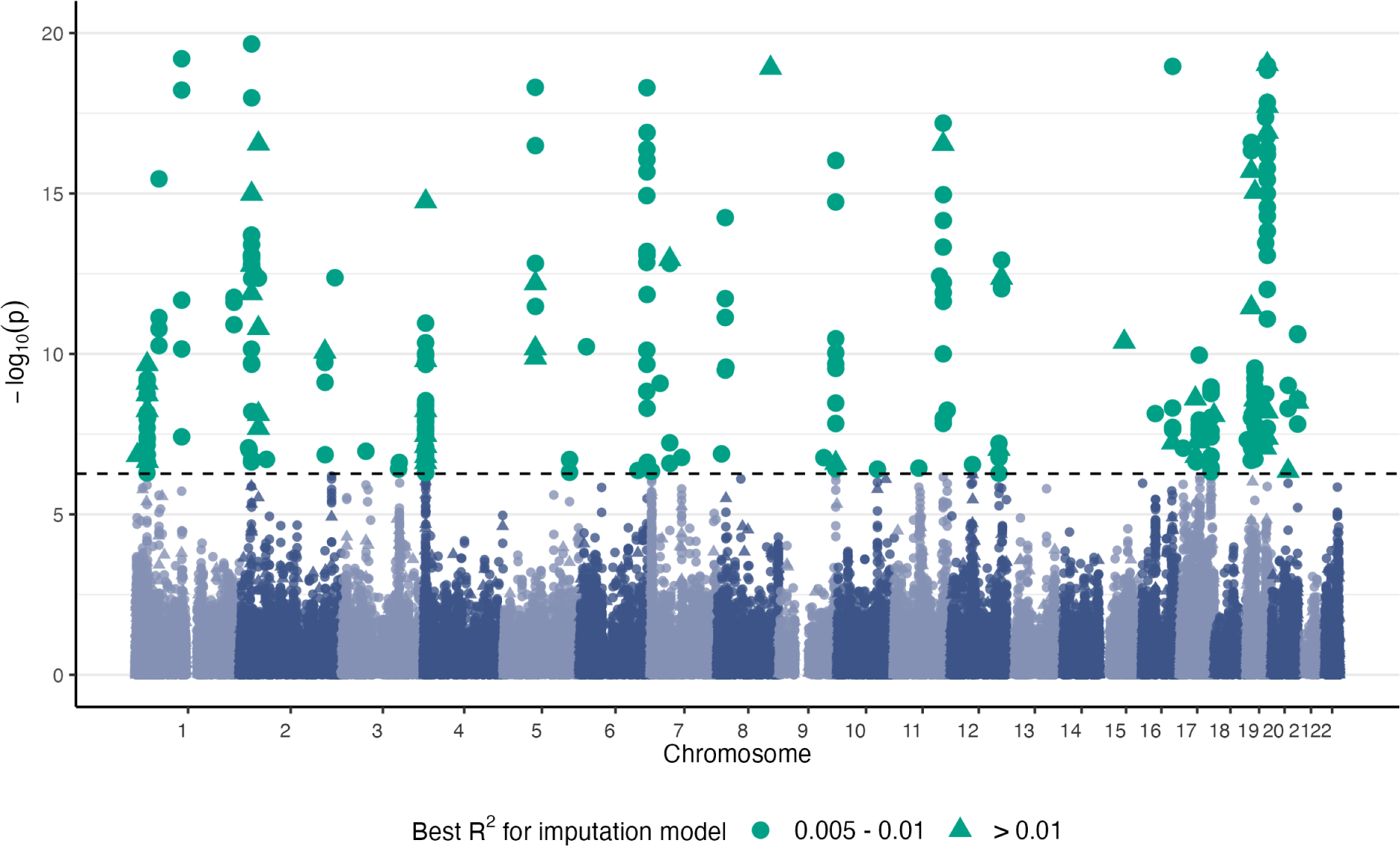
HCH Manhattan plot. CpG sites significantly associated with high cholesterol by MIMOSA at a Bonferroni-corrected p-value threshold 5.42 × 10^−7^.

We further explored the reasons for the superior performance of MIMOSA compared to Baselmans models. There are three key differences in the training of the MIMOSA and Baselmans models: 1) the significantly larger sample size for the MIMOSA training dataset compared to Baselmans; 2) MIMOSA combines five penalized regression models, Baselmans uses only LASSO; 3) the MIMOSA models are trained on SNPs within 1 Mb of the CpG sites, Baselmans models are trained on SNPs within 25 kb of the CpG sites. The majority of the improvement for MIMOSA vs. Baselmans is driven by the larger, summary-level, training data. We demonstrated through simulations that increasing the sample size of the training data massively increases the accuracy of DNAm prediction models (Supplementary Figure 3). The combination of penalized regression models does improve accuracy as well, though not nearly as drastically (Supplementary Figure 1). Lastly, through the examination of a subset of CpG sites in the test dataset, we determined that the larger *cis*-SNP window size for MIMOSA likely favors it over Baselmans, but only marginally so (Supplementary Figure 4). In summary, the ability to train on summary-level mQTLs and resultingly larger sample sizes drives the improvement of MIMOSA beyond the Baselmans method.

### 2.3. MIMOSA identifies more CpG site-trait associations

To assess the capability of MIMOSA to identify significant associations, we applied MIMOSA DNAm prediction models to the GWAS summary statistics of 28 complex traits and compared the results with those using the Baselmans models. The significant results are summarized in Supplementary Table 1. As discussed previously, MIMOSA assesses CpG sites with imputation *R*^2^ *>* 0.005 and positive correlation; however, we focused only on CpG sites with *R*^2^ *>* 0.01 and positive correlation here for a fair comparison. This slightly favored the competing method, as we used a more conservative threshold on MIMOSA. With this restriction, MIMOSA identified a total of 15,151 significant associations, a significant increase over the Baselmans method, which identified 7,680 (*p* = 1.0 × 10^−4^ per the Wilcoxon signed-rank test). MIMOSA further pinpointed 2,723 associations derived from CpG sites with 0.005 *< R*^2^ ≤ 0.01, bringing the overall association total to 17,874.

Since the two models have distinct sets of candidate CpG sites, we also compared MWAS results on the shared set of CpG sites for which both methods had imputation *R*^2^ *>* 0.01 and positive correlation. For these 55,131 candidate sites, MIMOSA again identified significantly more associations (10,408 vs. 6,098, *p* = 8.6 × 10^−5^). For the 6,098 associations identified by the Baselmans method, MIMOSA successfully identified 4,793 (78.6 %) under the Bonferroni correction and 5,654 (92.7 %) under a nominal p-value threshold *p <* 0.05.

Furthermore, MIMOSA effectively analyzed CpG sites with lower estimated heritability (0.005 ≤ *R*^2^ *<* 0.01). Still restricting to CpG sites with positive correlation, a total of 91,342 CpG sites had *R*^2^ *>* 0.005, of which 12,873 had 0.005 ≤ *R*^2^ *<* 0.01. For these CpG sites with lower heritability (0.005 ≤ *R*^2^ *<* 0.01), we found 2,723 significant CpG site-trait associations. For the remaining 78,469 CpG sites with *R*^2^ *>* 0.01, we identified 15,151 significant associations. This finding indicates that CpG sites with lower *R*^2^ (lower estimated heritability) are just as integral as those with higher *R*^2^, as the ratio of significant associations to CpG sites were very similar, 21.1% vs. 19.3%. We explored this further and examined the ratios of significant associations to CpG sites with *R*^2^ in bins of width 0.005 from (0.005, 0.845), where 0.845 is the maxmimum observed *R*^2^. We conducted a Kruskal-Wallis test to determine if these ratios could have come from different distributions; the resulting p-value was 0.49, which indicates that this was not the case. As a result, we concluded that lower heritability CpG sites are just as key as those with higher heritability. Notably, a similar finding has been reported for gene expression [8, 17]; researchers report that genes with low expression heritability have substantially larger causal effect sizes on complex traits [17].

### 2.4. MIMOSA determines putatively causal CpG sites for high cholesterol

We conducted an MWAS with MIMOSA for self-reported high cholesterol using GWAS summary statistics from 337,119 participants (41,296 cases) with European ancestry from the UK

Biobank. We identified 321 significantly associated CpG sites with at a Bonferroni-corrected threshold 5.47 × 10^−7^. For comparison, the Baselmans method found 165 significantly associated CpG sites, 128 of which were successfully identified by MIMOSA under the Bonferroni correction and 148 of which were identified by MIMOSA under the nominal p-value threshold 0.05. For the 321 CpG sites pinpointed by MIMOSA, we conducted sensitivity analysis with 2ScML [18], a new robust TWAS method for identifying causal effects in TWAS-style studies; this proceeded to indicate 146 of the associations were putatively causal.

We further conducted gene-set enrichment analysis with FGSEA [19] to link differential methylation of CpG sites along gene pathways to high cholesterol. Eight gene-sets were identified as significantly underenriched in methylated CpG sites in participants with high cholesterol by BH-adjusted p-values: scavenging by class F receptors, binding and uptake of ligands by scavenger receptors, scavenging by class H receptors, scavenging by class A receptors, chylomicron remodeling, plasma lipoprotein remodeling, regulation of toll-like receptors (TLRs) by endogenous ligands, and post-translational protein phosphorylation. There is prior biologic knowledge supporting links between the first seven of these pathways and high cholesterol.

To expand, the first four of the aforementioned pathways (scavenging by class F receptors, binding and uptake of ligands by scavenger receptors, scavenging by class H receptors, and scavenging by class A receptors) involve scavenger receptors, which are a set of receptors that can recognize molecular patterns associated with danger [20]. Specifically, these receptors are often involved in the uptake of modified low-density lipoproteins (LDLs) that encourage cellular accumulation of cholesterol [21, 22]. The next two (chylomicron remodeling and plasma lipoprotein remodeling) deal with remodeling lipoproteins. Chylomicrons are large, diffuse LDLs produced to facilitate the transport of dietary tryglycerides and cholesterol by binding to them, though they must be converted to chylomicron remnants in plasma to bind to LDL receptors on the surface of liver cells to remove cholesterol from circulation [23]. Lastly, endogenous ligands bind to receptors to produce physiological responses; damage-associated molecular patterns (DAMPs) fulfill this role for TLRs to regulate the innate immune system. Furthermore, DAMPs have been linked to the pathogenesis of atherosclerosis [24], in which plaques, including cholesterol, harden in the arteries. The finding of a potential role of post-translational protein phosphorylation for high cholesterol is rather novel, which warrants further investigation in the future. In a broader perspective, these results underscore the capacity of MIMOSA to effectively generate improved methylation prediction models, thereby boosting the results in downstream analyses. The consistency of most of our findings with existing literature provides a reassurance against the concerns of false positives.

### 2.5. Simulation studies confirm the superior performance of MIMOSA

We conducted a simulation study to confirm the superior performance of MIMOSA models. To save space, we describe the simulation results in the supplementary material, rather than the main body of the work. Briefly, in simulations based on a randomly selected CpG site, cg00110846, with varying values for methylation heritability 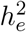, causal proportion of SNPs *p*_causal_, and phenotypic heritability 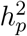, MIMOSA consistently outperforms the Baselmans method in both DNAm prediction accuracy and power to detect associations in MWAS (Supplementary Figure 2). Furthermore, we explored the impact of the sample size of the mQTL training dataset on DNAm prediction accuracy. Naturally, prediction accuracy, or *R*^2^, increases as the sample size increases. Additionally, for the true sample size of the GoDMC data, 27, 750, MIMOSA is able to perform nearly as well when trained with summary-level mQTLs (*R*^2^ = 0.093) vs. individual-level mQTLs (*R*^2^ = 0.094).

### 2.6. Data Resources

As the MIMOSA models are provided as a resource to the community, we wish to make clear how the resources may be used. We constructed a flow chart (Figure 5) for common scenarios in which one may want to use our resources. We anticipate that there are three such cases.

**Figure 5.**
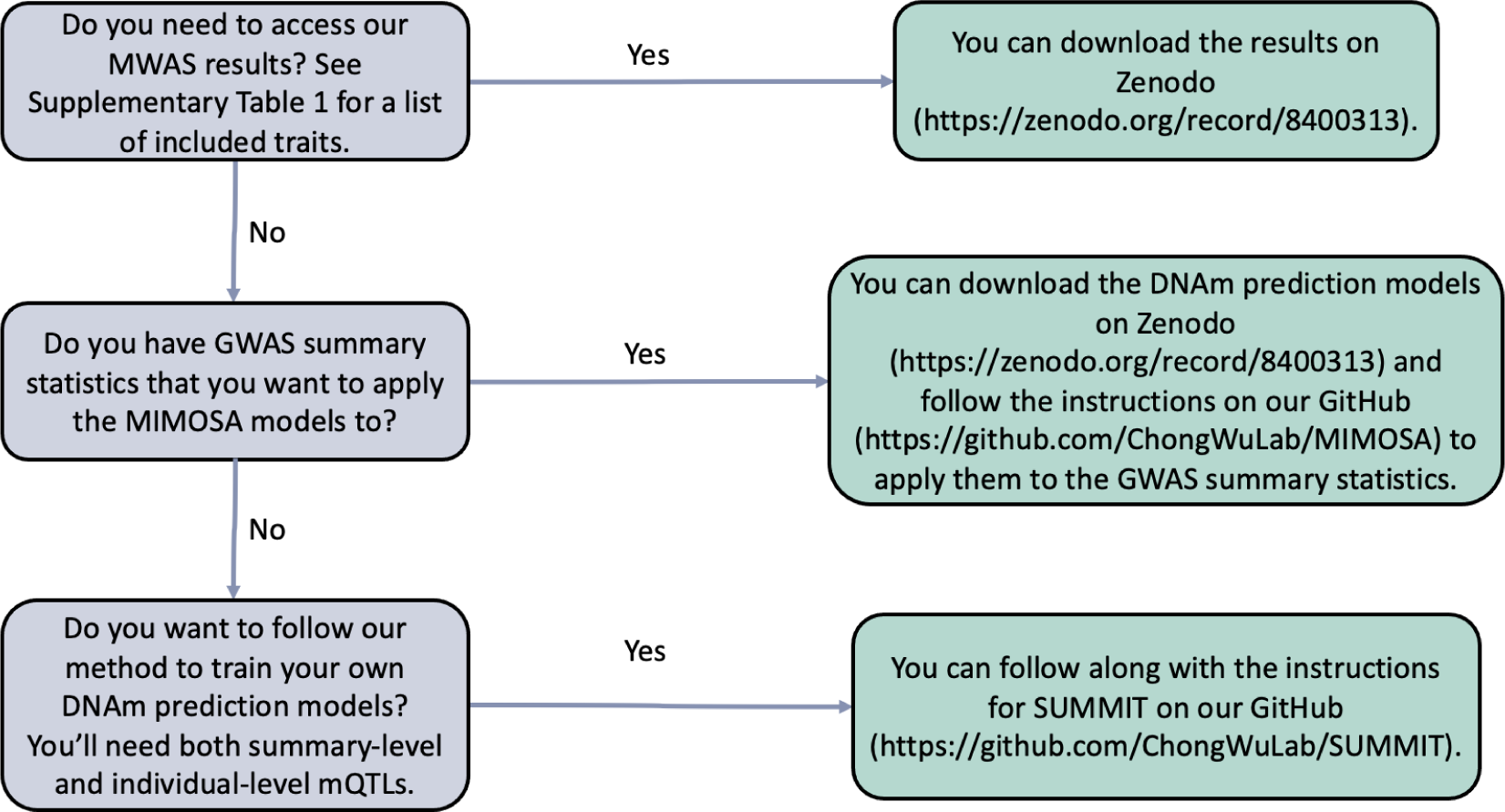
Flow chart for using MIMOSA.

First, we provide the full MWAS results for 28 GWAS traits on Zenodo https://zenodo.org/record/8400313. Simply download the *MIMOSA MWAS Results.zip* file, unzip the directory with whatever method is preferred, and the full set of MWAS results are available as text files.

Second, we provide the newly built DNA methylation prediction models (MIMOSA), which are also available on Zenodo https://zenodo.org/record/8400313. Download and unzip the *MIMOSA-Models.zip* file, and models for each of the CpG sites we considered will be available. Instructions for preparing GWAS summary statistics and running the MIMOSA models are available on our GitHub https://github.com/ChongWuLab/MIMOSA.

Third, one may have a different set of summary-level mQTLs, perhaps from another ancestry group or tissue, and wish to train their own set of DNAm prediction models. This can be accomplished by following the SUMMIT [8], for which instructions are also available on our GitHub https://github.com/ChongWuLab/SUMMIT.

## 3. Discussion

We built a comprehensive source of DNAm prediction models MIMOSA, which is created following the SUMMIT method [8] with large, summary-level mQTL datasets. MIMOSA notably improved imputation accuracy for the methylome and, through this, increased power to identify associations in MWAS by successfully leveraging the large size of the GoDMC [9] training data. We demonstrated through the analysis of GWAS results for complex traits that MIMOSA provided an appreciable step forward from previous MWAS models that were only able to be trained with individual-level mQTL data. MIMOSA increased accuracy in methylation imputation, identified more CpG site-trait associations, and was able to analyze CpG sites with lower heritability (i.e., imputation 0.005 ≤ *R*^2^ ≤ 0.01). Furthermore, as larger summary-level mQTL datasets are released, it is eminently possible to scale the MIMOSA models up by retraining them on the larger samples, further improving results.

MWAS can be viewed as one type of instrumental variable (IV) regression, which seeks causal relationships between exposures and outcomes using instrumental variables [25, 26]. From this perspective, the optimal instrument is the genetic prediction score combining information from multiple SNPs and providing the best prediction accuracy [27]. This explains why the MIMOSA models, which feature better prediction accuracy, increase the statistical power to identify (causal) associations in MWAS. On the other hand, it is important to note that causal interpretations are valid only when all genetic variants used to predict DNAm are valid IVs [5]. Horizontal pleiotropy is widespread [28] and violates instrumental variable assumptions. It is important to model the potential violation of valid IV assumptions, and we leave such interesting and exciting topics to future research.

The new MIMOSA resource is largely motivated by our previous work SUMMIT [8], where we build gene expression prediction models for TWAS using SUMMIT with summary-level eQTL data. While MIMOSA follows SUMMIT, the resource is equally important and widely needed by the community, as powerful DNA methylation imputation models such as those featured here will help us gain substantial insights into the etiology of complex diseases [29, 30].

We conclude by discussing the limitations of the current study. First, MIMOSA is built on mQTL data from whole blood from participants of European ancestry. Thus, the provided methylation imputation weights are appropriate for identifying CpG sites in whole blood for participants of European ancestry. With the eventual release of large, summary-level mQTL datasets for participants of other ancestry groups or using samples from other tissues, the MIMOSA method can be applied to train and tune new models appropriate for diverse cohorts. Fortunately, as blood cells play a key role in the immune system and are relevant to many phenotypes, this is a natural place to start. Second, the summary-level mQTL data used in MIMOSA are based on microarray technology (Illumina HumanMethylation BeadChips and Epic arrays), limiting our analysis to only 1.5% of the ∼28 million DNAm sites across the genome [31]. With the recent advancement in whole-genome bisulfite sequencing, we expect the percentage of analyzable CpG sites to grow substantially in the future. The framework can be equally applied once such summary-level datasets become available to the community. Additionally, while we cannot currently directly use individual-level mQTL data to train the MIMOSA models, such data could potentially be converted to the summary-level and combined with other summary-level data (e.g., GoDMC) through meta-analysis to further improve the prediction model performance. Additionally, we focused on cis-SNPs to build imputation models and did not take into consideration the effect of *trans*-mQTLs. Possibilities for future work include incorporating such *trans* information or leveraging methylation data generated from samples other than whole blood, similar to TWAS methods UTMOST [7] and MR-JTI [32]. We anticipate either direction will further improve the performance, though substantial additional work may be needed. For example, the effect size of trans-mQTLs is often weaker and is more likely to be impacted by horizontal pleiotropy [9]. Leveraging information generated from outside of the blood may also violate the valid IV assumptions because the used IVs may affect several tissues simultaneously. Finally, as discussed above, the causal argument of MWAS results (including using MIMOSA models) carry weight only when instrument variable assumptions hold. Therefore, careful interpretation of MWAS results is warranted and additional sensitivity analyses such as 2ScML [18] should be conducted.

## 4. Methods

We apply SUMMIT to build the MIMOSA models, a set of DNA methylation prediction models using summary-level mQTL data. For completeness, we briefly describe the methods here. A more detailed discussion may be found in Zhang et. al. [8].

### 4.1. Penalized regression model for methylation imputation

Consider the linear regression model for estimating DNA methylation levels shown below:

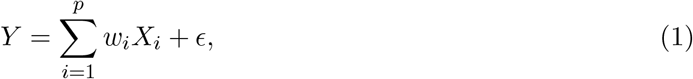

where *Y* is the *N* -dimensional vector of DNAm levels for a given CpG site, corrected for covariates such as age, sex, and genetic principal components. *X* = (*X*_1_′, …, *X*_*p*_′)′ is the standardized genotype matrix of *p cis*-SNPs around the CpG site. *w* = (*w*_1_, …, *w*_*p*_)′ is the mQTL effect size, and *ϵ* is mean-zero random noise.

We estimate the effect size *w* via five distinct penalized regressions, specifically LASSO [14], elastic net [10], the minimax concave penalty (MCP) [13], the smoothly clipped absolute deviation (SCAD) [12], and MNet [11]. Thus, we seek to minimize the objective function

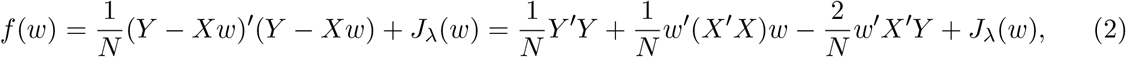

where *J*_*λ*_(*w*) is the primary penalty term. Then (2) contains 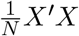, the linkage disequilibrium (LD) matrix for the SNPs, and 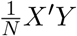, the standardized marginal effect size for SNPs (correlation between SNPs and DNA methylation levels). Denote the LD matrix with *R* and the standardized marginal effects with *r* = (*r*_1_, …, *r*_*p*_)′. We may also ignore 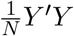, as it is not a function of *w*. Thus, the updated objective function is

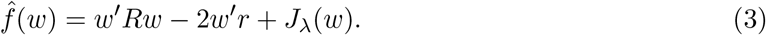

We estimate *r* with 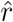, which comes from z-scores calculated from the summary-level mQTL database. We estimate *R* with 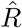, which comes from an LD reference panel (e.g., the 1000 Genomes Project [33]) used with a shrinkage estimator. Then the final objective function is approximated by

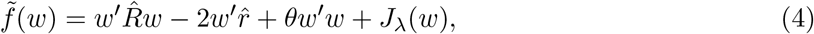

where *θw*′*w, θ* ≥ 0 is an additional *L*_2_ penalty designed to ensure a unique solution for 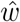. We include the additional penalty following [34], as we have estimated *X*′*X* using 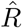 and *X*′*Y* using 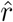, so the expression is no longer a penalized least squares problem and the solution would often be non-unique and unstable. The additional penalty term regularizes the expression. We then optimize 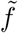.

The solution to the optimization, 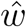, is found according to the coordinate descent algorithm [35], which is known to converge to a local minimum. The choice of *θ* is restricted to the set {0.1, 0.2, …, 0.9}, and a warm start is applied to generate a solution path for *λ*. These tuning parameters are chosen to maximize *R*^2^ in the individual-level tuning dataset, subcohort 1 of the FHS [15].

### 4.2. Methylation model training, tuning, and testing

The methylation imputation models were trained on summary-level mQTL data from GoDMC [9]. The GoDMC mQTL associations, simply the effect sizes of SNPs on DNAm, are based on a sample size of 27,750 participants of European ancestry from 36 independent studies, and 102,965,711 candidate *cis*-mQTLs were identified (*p <* 1 × 10^−5^, ±1 Mb from DNAm site) [9]. DNAm was profiled in whole blood with Illumina HumanMethylation450 BeadChips or EPIC arrays in at minimum 100 European ancestry participants. The DNAm data were normalized and quality-controlled (described further in [9]) and adjusted for age, sex, predicted cell counts, predicted smoking, genetic PCs, and nongenetic PCs. In the case of related participants, each of the same steps was performed in addition to fitting the genetic kinship matrix. Finally, residuals from the previous analyses were centered and standardized to have mean 0 and variance 1. Additionally, SNPs located in the MHC region (26 MB - 34 MB on chromosome 6) were removed due to its known complex LD structure [36].

To select tuning parameters for the model, such as the previously mentioned *λ*, and *θ*, we employ DNA methylation and genotype data from the Framingham Heart Study subcohort 1 (*n* = 797) [15]. DNAm was profiled using the Illumina HumanMethylation450 BeadChip. We focused on unrelated individuals of European ancestry. The DNAm data were quality controlled and normalized (as described in [4]) and adjusted for age, sex, six cell-type composition variables (CD8 T cells, CD4 T cells, NK cells, B cells, monocytes and granulocytes) using the R package minfi [37], and the top ten principal components (PCs) from the genotype data. The best tuning parameters were selected by examining *R*^2^, the proportion of variance of the DNAm that can be explained by the DNAm prediction models.

To validate the model choices, we employed an independent subset of data from the Framingham Heart Study subcohort 2 [15], with no overlap from the previous tuning data (*n* = 798). The validation data was processed in accordance with the tuning data, and we selected models for methylation imputation with *R*^2^ *>* 0.005.

### 4.3. Applying MIMOSA to GWAS

The MIMOSA models can be applied to both individual- and summary-level GWAS data. For individual-level GWAS data, use a generalized linear regression model:

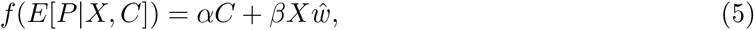

where *P* is the phenotype for the GWAS, *X* is the genotype data (and thus 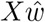 is the imputed methylation), *C* is the matrix of covariates, and *f* is a chosen link function. This expression is used to test null hypothesis *H*_0_ : *β* = 0, i.e., does methylation affect the phenotype.

For summary-level GWAS data, apply a burden-style test:

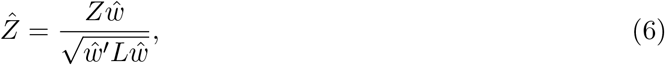

where *Z* is a vector of z-scores for all analyzed SNPs, 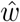 is the estimated effect size, and *L* is the LD matrix for those SNPs (following SUMMIT, *L* is estimated by a population LD reference panel such as from the 1000 Genomes project [33]).

While the above discussion on applying the MIMOSA models assumes only a single DNAm prediction model, MIMOSA can include up to five. When multiple methods build satisfactory prediction models for a single CpG site (DNAm prediction *R*^2^ *>* 0.005, positive correlation), following SUMMIT, we conduct an association test as in the above procedure. The results are consolidated for the CpG site with the Cauchy combination test [38]. In detail, assuming *K* satisfactory models, the test statistic

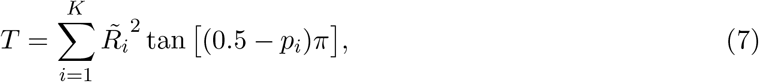

approximately follows the standard Cauchy distribution, and 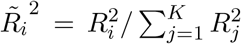 and *p* is the p-value for the *i*th model. The consolidated p-value is found with *p* = 0.5 − arctan(*T*)*/π*. Notably, the test statistic *T* is weighted by the DNAm prediction accuracy, so better fitting models are more impactful in the consolidated p-value.

### 4.4. Comparison with existing methods

To demonstrate efficacy and utility, we compared MIMOSA with the DNA methylation imputation model weights generated by Baselmans et al. [2]. These weights were built from individual-level, whole-blood methylation data from the BIOS Consortium [16] using 4,008 samples measured via Illumina 450k arrays. The BIOS Consortium provides genetic, methylome, transcriptome, and phenotypic data on individuals from six Dutch biobanks. For each CpG site with an mQTL (151,729), a DNAm imputation model was created using LASSO penalized regression [14] with all nearby (within 2.5 × 10^4^ bp, this is discussed further below) SNPs. Then, with the imputation models, summary-level GWAS for a given phenotype, and LD reference from the 1000 Genomes Project [33], a burden-style test based on test-statistic 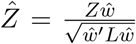 is performed, exactly as with MIMOSA.

For the comparison, we first examined methylation prediction accuracy on the aforementioned validation set of data from the FHS, conducted by examining *R*^2^ and correlation at each CpG site for which both methods possess weights. We tested the differences in accuracy between the two methods with the one-sided Wilcoxon signed-rank test, which compares the locations of matched samples to determine if their population means differ. We further compared the methods by analyzing GWAS summary-level data for 28 complex traits. For each method and trait, we used the Bonferroni correction to determine significance level thresholds. In an effort to make an unbiased comparison, we further compared the methods for their shared CpG site set and used identical Bonferroni-corrected significance cutoffs. We then tested the difference in the number of significant CpG sites for the two methods with the Wilcoxon signed-rank test.

We note that while both the Baselmans models and the MIMOSA models were trained on *cis*-SNPs, these are defined differently for the two sets of training data. For MIMOSA, the GoDMC summary-level mQTLs include all SNPs within 1 Mb (1 × 10^6^ bp) of the CpG site. For Baselmans, the BIOS Consortium mQTLs include all SNPs within 25 kb (2.5 × 10^4^ bp) of the CpG site. To ensure that this difference in *cis*-SNP window size did not drive the increased performance of MIMOSA, we compared the correlation between predicted DNAm levels and actual DNAm levels and the accuracy of DNAm prediction models for MIMOSA with the typical *cis*-SNP window size 1 Mb to MIMOSA with the smaller *cis*-SNP window size 25 kb (Supplementary Figure 4) on 10,000 randomly selected CpG sites. The models built with the larger *cis*-SNP window size yielded comparable results as that using the smaller window size (25 kb). In more detail, for the 10,000 compared CpG sites, the larger window size models had higher correlation in 4,620, lower correlation in 4,453, and equal correlation in the remaining 927. For prediction accuracy *R*^2^, the larger window size was better for 4,600 CpG sites, worse for 4,474, and equal for 926. Given the sample size of GoDMC data is large (*n* = 27, 750) and we hope to consider all *cis*-SNP when constructing methylation prediction models, we used SNPs within 1 Mb as our predictors. We leave interesting topics such as the impact of window size to future research.

### 4.5. Application to HCH GWAS

We applied MIMOSA to a set of GWAS summary data for self-reported high cholesterol from the UK Biobank to identify associated CpG sites. The data consists of 337,119 participants of European ancestry, and more details about the data are available on the UK Biobank’s website (https://biobank.ndph.ox.ac.uk/ukb/field.cgi?id=20002). Putatively causal CpG sites were identified by sensitivity analysis with 2ScML [18]; these results are available in the supplementary data. We then conducted gene-set enrichment analysis with FGSEA [19] to associate differential methylation along gene pathways with high cholesterol.

### 4.6. Simulation design

We conducted a simulation study to accomplish the following goals. First, assess the accuracy of MIMOSA DNAm imputation models and their corresponding power to detect associations with traits. Second, evaluate the impact of mQTL reference panel size. Third, ensure MIMOSA can achieve similar results when using a summary-level vs. individual level mQTL reference panel.

To accomplish these, we employed data from the UK Biobank (application number 48240). Genotype data were taken from the 27,750 (matching the sample size of the mQTL data from GoDMC) randomly chosen and unrelated individuals of White British descent for training the DNAm imputation models. Tuning data were drawn from an independent set of 797 (matching the sample size of the tuning data from the FHS) White British individuals, and testing data were drawn from a further 10,000 unrelated White British subjects. We randomly selected the CpG site cg00110846 on chromosome 7 for simulations.

DNAm levels and phenotype values were simulated according to equations *M* = *Xw* + *ϵ*_*m*_ and *P* = *βM* + *ϵ*_*p*_ respectively. Here, *X* is the standardized genotype matrix, *w* the vector of effect sizes, 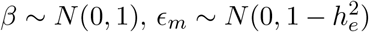, and 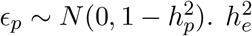 represents the DNAm heritability (proportion of variance of DNAm explained by SNPs) and 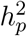 represents phenotype heritability (proportion of variance of phenotype explained by SNPs). Let *p*_causal_ be the proportion of causal SNPs in the DNAm model, then we simulated SNP effect size *w* ∼ *N* (0, 1). *l* SNPs were then selected to retain their weights, and the remaining weights set equal to zero in order to ensure that *p*_causal_ of the SNPs were truly causal. Effect sizes *w* and *β* were then re-scaled according to 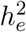 and 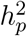.

To evaluate the performance of MIMOSA, we performed an association scan on the entirety of the simulated training data (*M, X*). Summary-level data were computed (Z-scores) by a linear regression. In addition to models built from summary-level data, individual-level data with disparate sample sizes (250, 500, 1000, 10000, 27750) were employed to build models; these scenarios allowed us to confirm that the quality (measured by *R*^2^) of DNAm imputation models increases with sample size and that MIMOSA can recapitulate individual-level results using summary-level data (where *N* = 27, 750) only. MIMOSA models were compared with the Baselmans method, where data of 4,008 individual-level samples from the BIOS Consortium were used to build models [2]. In accordance with real data, primary comparisons between the two methods were made using summary-level mQTLs from 27,750 samples for MIMOSA, individual-level mQTLs from 4,008 samples for Baselmans, and corresponding LD reference panels for both methods.

We attempted to cover a wide range of scenarios in the simulation study by varying the DNAm heritability 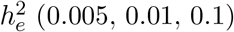, the phenotype heritability (0.1, 0.2), and the proportion of causal SNPs *p*_causal_ (0.01, 0.1, 0.2). Each simulation scenario was repeated 1,000 iterations, and statistical power was determined by the proportion of those iterations that resulted in a p-value less than the genome-wide significance threshold 1.39 × 10^−6^.

## Supporting information

Supplementary Results

## Code availability statement

Code to use the MIMOSA models in MWAS is available on GitHub(https://github.com/ChongWuLab/MIMOSA. Code and data to reproduce the results given in this study are available on Zenodo https://zenodo.org/record/8400313.

## Data availability

The mQTL summary data are available at http://mqtldb.godmc.org.uk/downloads. The datasets of FHS Offspring Cohort are publicly available via dbGaP (www.ncbi.nlm.nih.gov/gap): dbGaP Study Accession: phs000342 and phs000724. The UK Biobank is an open-access resource but requires registration, available at https://www.ukbiobank.ac.uk/researchers/. The Basel-mans models can be downloaded from http://bbmri.researchlumc.nl/atlas/#data. The 1000 Genomes Project data can be downloaded from https://www.internationalgenome.org/data. The genetic distance data for 1000 Genomes Project can be downloaded from https://github.com/joepickrell/1000-genomes-genetic-maps. The MIMOSA models are available at Zenodo https://zenodo.org/record/8400313. Real data results are available at Zenodo as well https://zenodo.org/record/8400313, where they can be easily downloaded.

## Acknowledgements

The content in this study is solely the responsibility of the authors and does not necessarily represent the official views of the National Institutes of Health. This study was conducted using the UK Biobank recourse under Application Number 48240 (https://www.ukbiobank.ac.uk/researchers/). We thank Dr. Jihwan Ha at University of Hawaii Cancer Center for his help for this study. The authors would like to thank all of the individuals for their participation in the GWASs and UK Biobank and all the researchers, clinicians, technicians and administrative staff for their contribution to the studies and for making their GWAS summary results publicly available.

