## Supplementary Results for "MIMOSA: A resource consisting of improved methylome imputation models increases power to identify DNA methylation-phenotype associations"

### 1 Simulation Results

We conducted a simulation study to confirm that MIMOSA indeed outperforms the existing Baselmans models in DNAm prediction and improves MWAS power. Initially, we evaluated the accuracy of the DNAm prediction models created by MIMOSA and Baselmans, along with the corresponding statistical power. We then assessed the impact of sample size on DNAm prediction performance and revealed that MIMOSA can effectively replicate the information of individual-level DNAm reference panels using summary-level information.

Supplementary Figure 2 conveys that MIMOSA outperforms the Baselmans models in terms of DNAm prediction model accuracy on a set of candidate values for proportion of causal SNPs ($\mathrm{p}_{\mathrm{causal}}$) and DNAm heritability ($h_e^2$). As an example, let $\mathrm{p}_{\mathrm{causal}} = 0.1$ and $h_e^2 = 0.01$; the median prediction $R^2$ was 0.68% for MIMOSA and 0.17% for Baselmans. The increased accuracy resulted in a corresponding increase in power for the downstream association studies (Supplementary Figure 2b). Again, for $\mathrm{p}_{\mathrm{causal}} = 0.1$, $h_e^2 = 0.01$, and now phenotypic heritability $h_p^2 = 0.2$, the power was

determined to be 0.993 for MIMOSA and 0.748 for Baselmans. This is mainly because MIMOSA was based on summary-level mQTL data with a much larger sample size (N = 27,750) than that of the Baselmans models, which were based on individual-level reference dataset with a comparatively small sample size ($N = 4,008$).

Notably, MIMOSA illustrated the ability to successfully predict DNAm levels in the low heritability setting ($h_e^2 = 0.005$). According to Supplementary Figure 2, the average imputation $R^2$ for MIMOSA is much closer to 0.5% than for Baselmans, regardless of the choice of $\mathrm{p}_{\mathrm{causal}}$ when $h_e^2 = 0.005$. For example, when $\mathrm{p}_{\mathrm{causal}} = 0.1$, prediction $R^2$ for MIMOSA was 0.29% vs 0.05% for Baselmans. Since we could predict DNAm levels in this setting reasonably well, we proceeded to include such CpG sites with $0.005 < R^2 \leq 0.01$ in our analysis with confidence.

We next explored the impact of sample size of the DNAm reference panel (Supplementary Figure 3). Naturally, imputation $R^2$ increased with sample size. To demonstrate, for DNAm heritability $h_e^2 = 0.1$, the average imputation $R^2$ increased from 3.30% to 9.44% when the sample size increased from 250 to 27,750. This result emphasized the importance of using larger reference panels. Furthermore, the imputation models increased in stability (lower variance) with increased sample size. Additionally, MIMOSA produced an average imputation $R^2 = 9.35\%$ when applied to summary-level mQTLs, very close to the average $R^2 = 9.44\%$ when the individual-level data were available, which validated that MIMOSA can faithfully recapitulate the individual-level information from summary-level data.

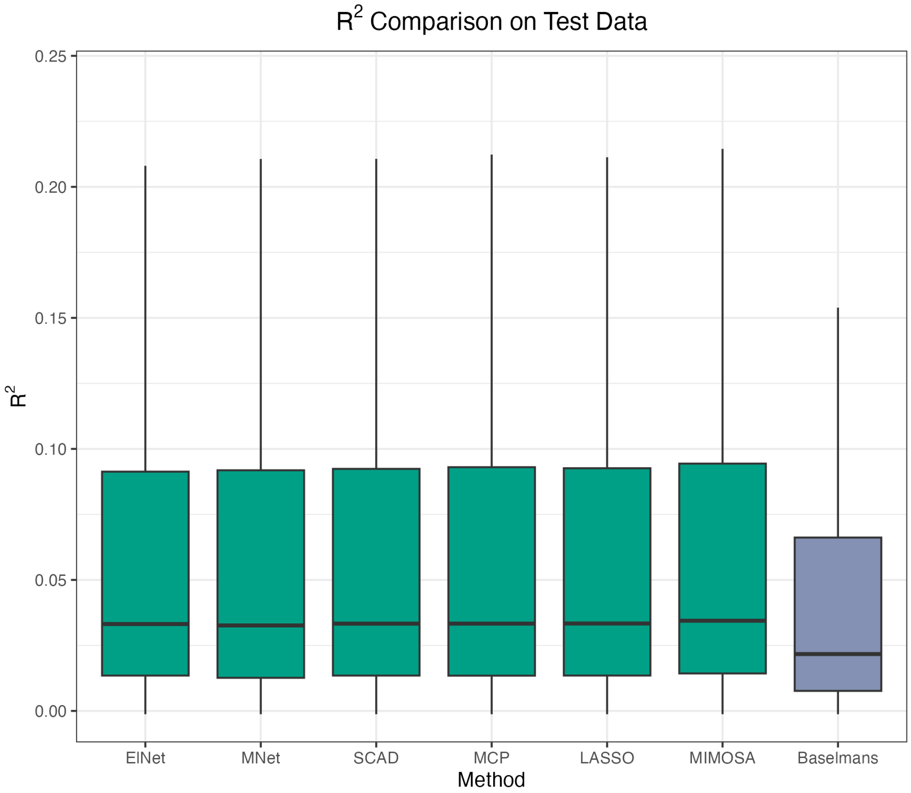


Figure 1: $R^2$ **comparison on test data for five penalized regression methods in MIMOSA and Baselmans.** A comparison of DNAm prediction accuracy for each of the five penalized regression methods as well as MIMOSA (the best of the five) and Baselmans.

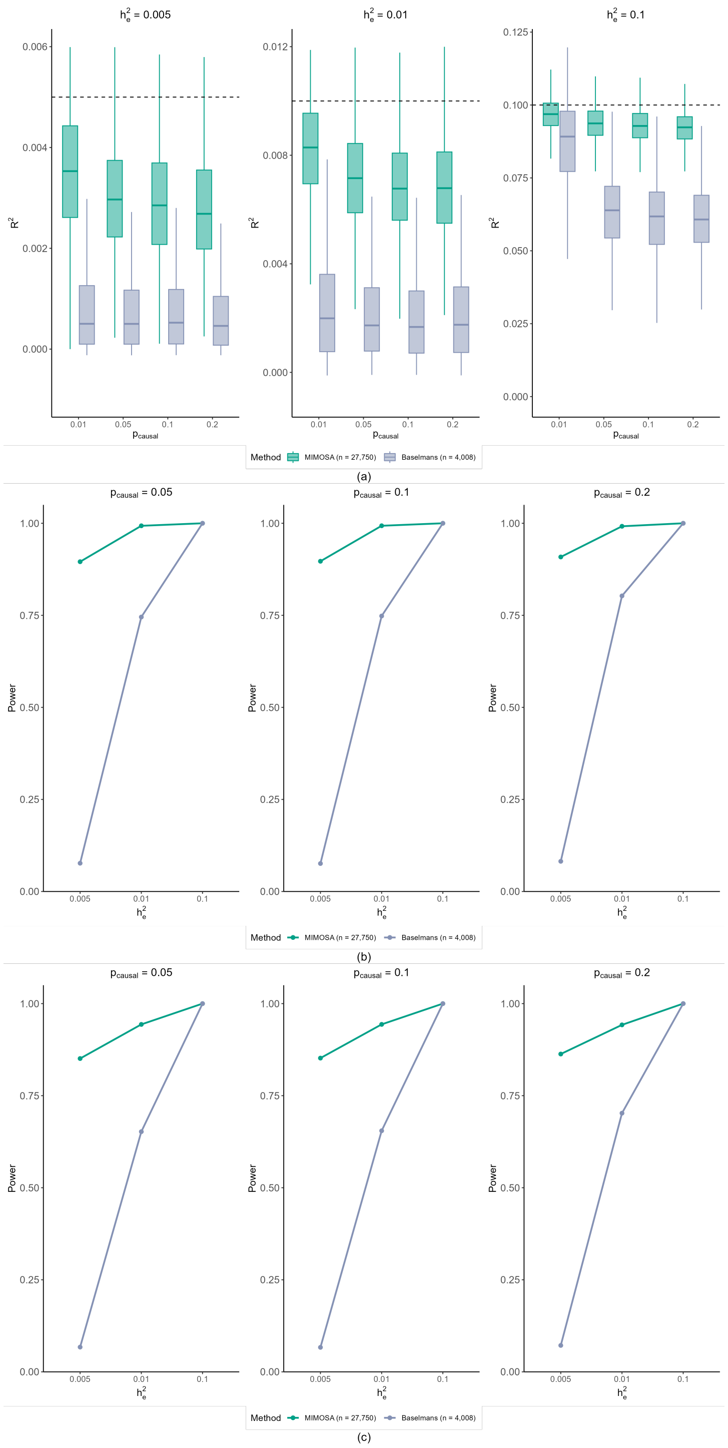


Figure 2: **Simulation results with CpG site cg00110846.** a) A comparison of DNAm prediction accuracy in simulations for a variety of settings for DNAm heritability ($h_e^2$) and percentage of causal SNPs ($p_{\text{causal}}$). b) A comparison of empirical power with different settings for DNAm heritability ($h_e^2$) and percentage of causal SNPs ($p_{\text{causal}}$) with phenotype heritability ($h_p^2$) 0.2. c) A comparison of empirical power with different settings for DNAm heritability ($h_e^2$) and percentage of causal SNPs ($p_{\text{causal}}$) with phenotype heritability ($h_p^2$) 0.1. Empirical power was determined by the proportion of those iterations that resulted in a p-value less than the genome-wide significance threshold $1.39 \times 10^{-6}$.

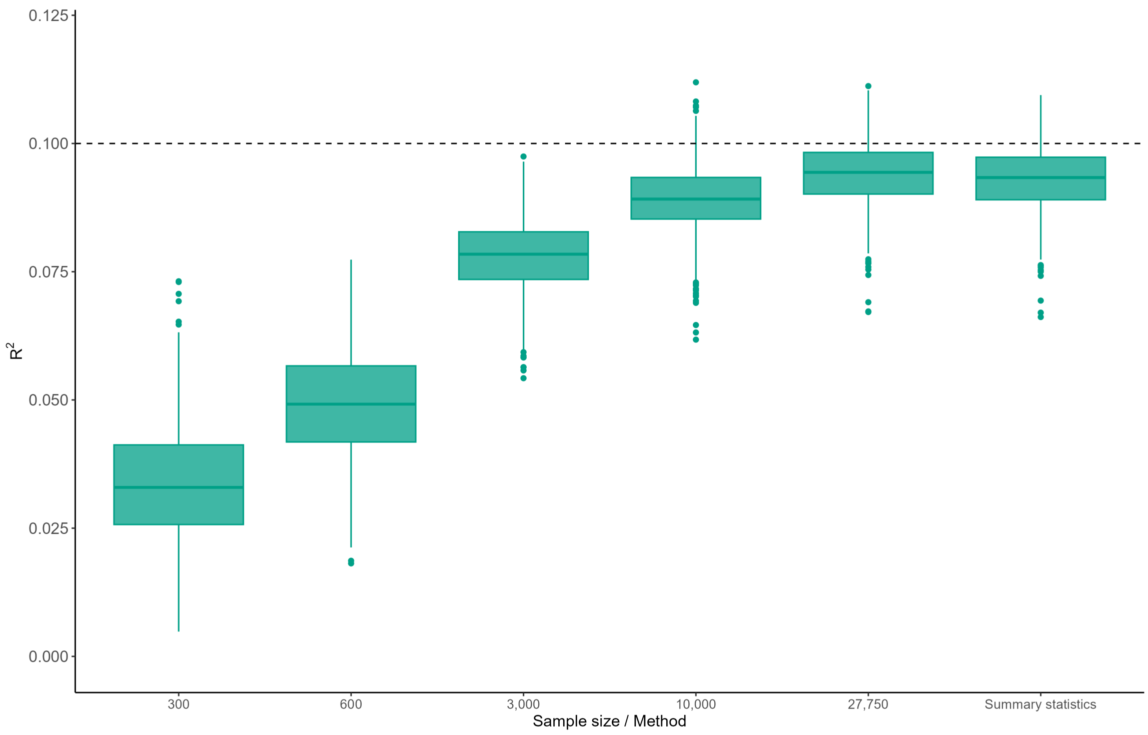


Figure 3: **Simulation results for MIMOSA DNAm prediction accuracy when varying sample size.** A comparison of DNAm prediction accuracy in MIMOSA models trained on different sample sizes of individual-level mQTL datasets and one summary-level mQTL dataset with sample size $N = 27,750$.

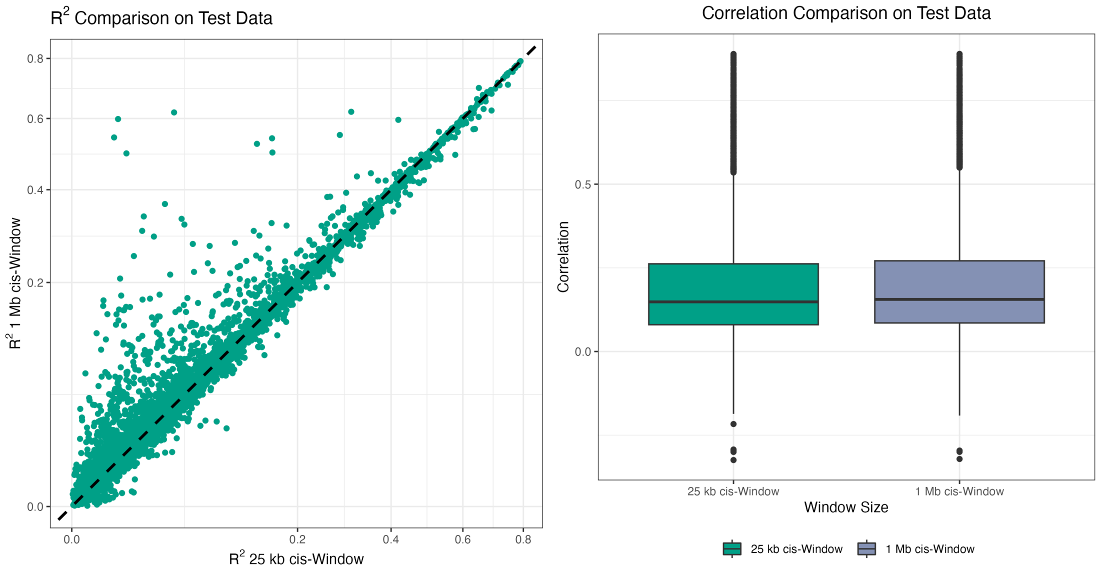


Figure 4: $R^2$ **and correlation comparison on a subset of test data for different** ***cis*****-SNP window size.**
